# Contributions of immune cell biomarkers to explaining differences in mortality risk by sex in the Health and Retirement Study

**DOI:** 10.64898/2026.05.27.26354256

**Authors:** Margaret Yin, Vy Kim Nguyen, Aparna Nathan, Chirag J Patel

## Abstract

**Background:** It is well-established that males have a higher mortality risk than females. Immune cells and their function are known to undergo characteristic changes during aging, and immune cells are known to have sex differences. Immune cells and their function have been linked to mortality risk, but no studies have investigated to what degree, if at all, Immune Cell Biomarkers (ICBs) contribute to the known differences in mortality risk by sex.

**Methods:** Using participant data from the Health and Retirement Study (n = 8,822), we applied multivariable linear regressions adjusting for age, cytomegalovirus (CMV) serostatus, sex, and race/ethnicity to identify differences by sex in 48 immune cell biomarker (“ICB”, e.g. T cells, B cells, Monocytes, etc.) percentages and counts (measured in 2016). We studied how the associations between ICBs and mortality risk differ by sex using stratified Cox Proportional Hazard (CPH) models. We estimated how inclusion of sex explained the relationship between ICBs and all-cause mortality, and conversely, how inclusion of individual and all ICBs combined explain the relationship between sex and all-cause mortality using multivariable modeling approaches.

**Results:** Differences in ICBs by sex range between 2-38% (39/48 statistically significant). 9 ICBs were significantly associated with mortality risk in the entire sample. While different ICBs were significantly associated with mortality risk in the stratified analyses, particularly with respect to monocyte, B cell, and NK cell populations, adjusting for sex modestly influenced the hazard ratios of the ICBs (sex: 8 ICBs, percent change <5.4%). Furthermore, individual and cumulative contributions of ICBs in explaining the differences in mortality risk by sex were not significant.

**Conclusion:** ICBs vary by sex in the older population; however, their role in explaining mortality risk by sex are modest.

## INTRODUCTION

All-cause mortality is the ultimate health outcome, with males having consistently higher risk for death around the world (1,2). Despite literature on mortality differences between males and females tentatively ascribed to sociological and biological factors that are potentially conserved (3–8), there are still many unanswered questions regarding which factors are most relevant to these mortality differences and how they may uniquely contribute to such differences.

It is possible to study how biological factors, such as immune system changes, may explain mortality disparities between males and females as they age. “Immunosenescence” is a phenomenon that describes characteristic changes in the immune system associated with aging, and it is often associated via changes in immune cell biomarkers (ICBs) (9,10) , such as immune cell counts and proportions. “Inflammaging” is a hallmark aging process and key feature of immunosenesence. This process describes a state of chronic, low-grade inflammation associated with increased expression of upregulated Interleukin (IL)-1, IL-6, IL-8, IL-13, IL-18 and Tumor Necrosis Factor (TNF), among other pro-inflammatory factors highly expressed by senescent cell (10,11). Additionally, thymic involution, or the degradation of thymic tissue, is also strongly implicated in immunosenescence, particularly regarding counts and proportions of T-cell populations; with aging, there is a general decrease in naïve and increase in late-differentiated memory T cells (10,12–14). Similarly, there is also a decrease in naïve B cell and increase in memory B cells associated with aging, as well as general decrease in B cell production (11,15).

Both at baseline and during aging, the immune system exhibits differences by sex. Generally, females are known to have stronger innate and adaptive immune responses and also show greater susceptibility to auto-immune and inflammatory diseases, especially with age (16,17). Regarding specific ICB differences, males are known to have higher counts of natural killer (NK) cells responsible for innate immunity as well as higher counts of CD8+ T cells and regulatory T cells. There is a higher ratio of CD4/CD8 Tcells, CD4+ T cell counts, and B cell counts in females (18). Sex differences in immune cell counts and inflammatory signaling have shown to be exacerbated with aging (19); notably, while inflammaging occurs in both sexes, it is more pronounced in males (16)(.

Do these ICBs – which are known to differ by sex and critically affected by aging – explain the sex differences mortality risk? To date, while previous studies have found relationships between ICBs, especially T-cell related counts and proportions, and risk for death (19,20), no studies have investigated the differences in how ICBs predict all-cause mortality risk by sex. Here, we analyze participant data from the Health and Retirement Study, a representative longitudinal cohort study replete with biomarkers of aging, including ICBs such as immune cell counts and proportions, to test whether they explain observed sex-related differences in mortality.

## METHODS

The HRS (Health and Retirement Study) is sponsored by the National Institute on Aging (grant number NIA U01AG009740) and is conducted by the University of Michigan (21). The HRS is a nationally representative longitudinal survey study of adults over 50 years of age in the United States. Funded by the National Institute of Aging, the HRS began in 1992 with additional interviews roughly every two years. The steady-state design replenishes the sample every six years with a younger cohort of older adults. The HRS sample is selected using a multi-stage area probability sample design. Black and Hispanic populations are oversampled at roughly twice the rate of White populations, and oversampling is adjusted for in the provided sampling weights. The study evaluates a breadth of variables on aging including economic status, family structure, expectations, biological measurements, and more (22).

We analyzed immune cell biomarkers (ICBs) located in the Flow Cytometry data file (23). The Flow Cytometry data file contains assay results from the whole blood samples collected in the 2016 wave for the Venous Blood Study (VBS). The flow cytometry data included characterizations of 33 different subsets of T, B, monocytes, and NK cells. Counts and percentages were often both given, and percentages were taken out of the parent population (e.g. CD16+ monocytes were defined as a percentage of total monocytes). All flow cytometry measurements were performed on an LSRII flow cytometer or a Fortessa X20 instrument (BD Biosciences, San Diego, CA). While these cell counts and percentages do not capture functional changes within individual cell subtypes, these are biomarkers that are reflective of broader changes in immune function and easily attainable from blood samples. Both cell percentages and counts were included as the two measurements may confer different information about composition of the immune system. To provide additional insight into functional changes in immune function, cytokine (IL10, IL6, IL-1RA, TNFR-1, and TGF-Beta) measurements were obtained from the VBS data file. Further details about flow cytometry are outlined in (24) and the documentation for the VBS data file. Cytomegalovirus (CMV) immunoglobin G (IgG) antibody levels were taken from the 2016 VBS (25).

The RAND HRS Longitudinal File is a harmonized data product created by the RAND Corporation. We used the RAND HRS Longitudinal File 1992-2018 to access age, mortality status, and sex variables (26–28).^1^ [^1^The RAND HRS Longitudinal File is an easy-to-use dataset based on the HRS core data. This file was developed at RAND with funding from the National Institute on Aging and the Social Security Administration.]

We used race/ethnicity variables and weights from the HRS Tracker File 2020 (29). The Tracker File contains one record for every person who was ever eligible to be interviewed in any wave.

This dataset has separate categories for Mexican Hispanic and Non-Mexican groups, but due to small sample size, we ultimately recoded participants into three racial/ethnic categories: Non-Hispanic White, Non-Hispanic Black, and Mexican and non-Mexican Hispanic. Survey weights for the 2016 Blood-Based Biomarker study were used because flow cytometry results were from samples collected through the blood biomarker study. We excluded any individuals with missing mortality status or racial/ethnic group information in re-coded variables as well as those with missing weights, which are to be excluded from analysis due to illegibility criteria determined by the HRS (n = 1098).

### Statistical Analysis

We performed all analyses using R. We applied the survey weights from the Blood Biomarker 2016 dataset to all statistical models to account for the HRS sample design and allow for results to be generalizable to the retired United States population. We used functions from the survey() R package (30).

#### Assessing sex differences in immunological biomarkers (ICBs)

We used multivariable regression models to assess the differences in immunological biomarkers by sex. We used a series of survey-weighted linear models with log10 transformed ICBs as the outcome variable and sex (categorical), race/ethnicity (categorical), age (continuous), and CMV IgG antibody levels (continuous) as covariates (**Figure 1**, Model 1). The reference group for sex was the male group. Age, measured in months, was calculated using the reported date of birth and the date of the 2016 interview at which the samples used for flow cytometry were collected. We controlled for race as previous studies on HRS data have shown that racial/ethnic groups had differences in aging-related immune markers (31,32). We controlled for CMV as previous literature has established that exposure to CMV infections affects distributions of ICBs (33). CMV seroprevalence was measured in the VBS using IgG antibodies to CMV in serum using the Roche e411 immunoassay analyzer (Roche Diagnostics Corporation, Indianapolis, IN) and assayed. The regression coefficient for sex was converted into percent differences ([10^coefficient^ – 1] * 100) for ease of interpretation. To identify significant differences but maintain a lower false positive rate, we applied the false-discovery rate (FDR) method to the p-values from the model outputs.

**Figure 1:**
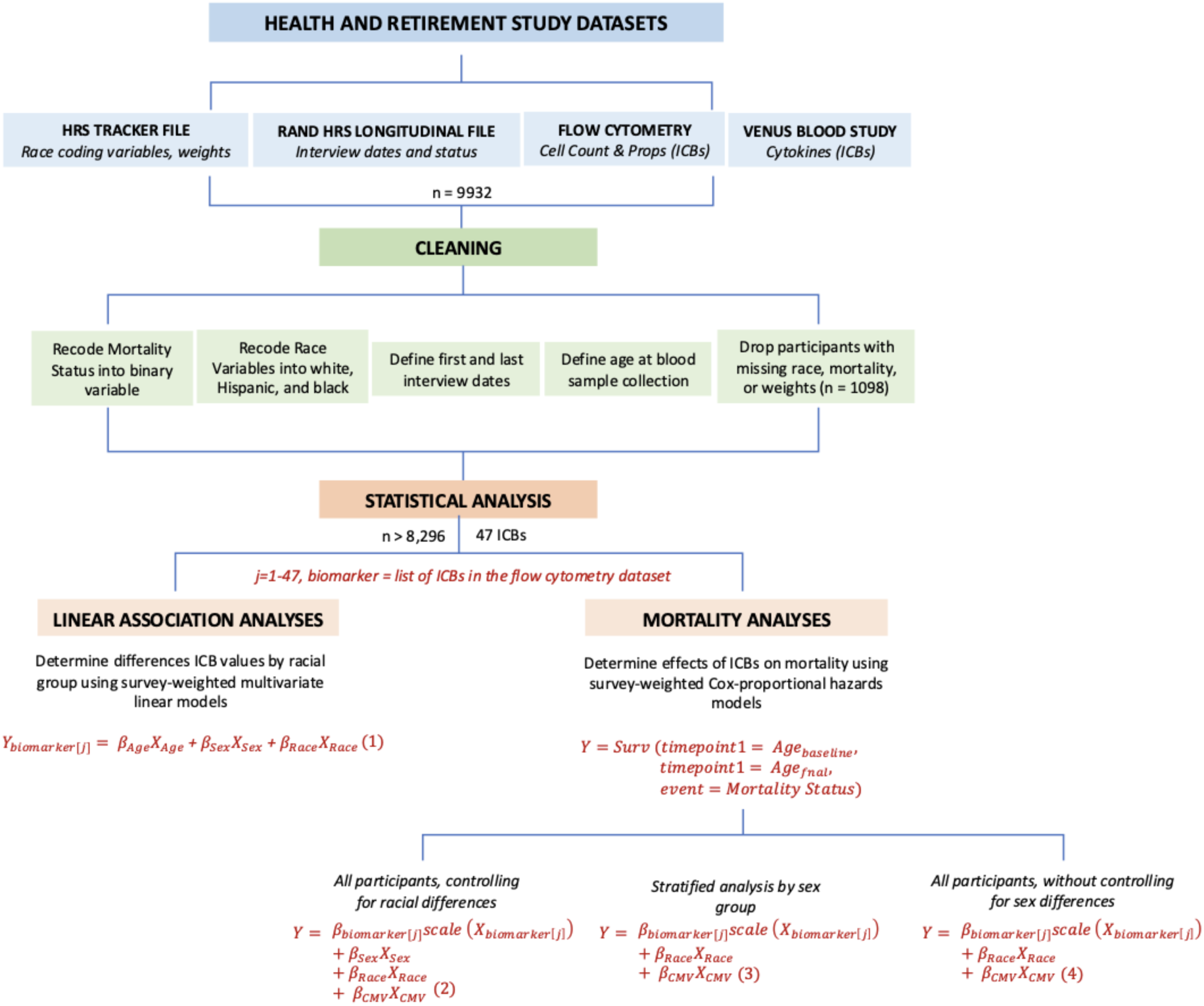
Methods Schematic. Variables from three HRS datasets (HRS Tracker File 2020, RAND HRS Longitudinal File 1992-2018, Flow Cytometry 2016) were merged into one dataset, which was wrangled and cleaned for analyses. Statistical analysis using this dataset included a linear association model (Model 1) and mortality analyses (Models 2, 3, and 4).

#### Associating ICBs with all-cause mortality

We then investigated how immunological biomarkers were associated with all-cause mortality. We first assessed how sex is correlated with all-cause mortality risk by applying survey-weighted Cox Proportional Hazard (CPH) models. For each immunological biomarker, we applied a CPH model with sex, race/ethnicity, and CMV as covariates (**Figure 1**, Model 2). Additionally, we also analyzed sex stratified associations (**Figure 1**, Model 3).

We used the time points method to accommodate the wave design of the HRS in which participants enter and exit in different waves. Using the birthdate and interview date, we calculated the age at the first baseline interview (timepoint 1) and the age at death or the most recent interview in 2018 (timepoint 2). We used the interview status (i.e. mortality status) from 2018, which is the most recent data available at the time of analysis.

Participants marked alive or presumed alive were categorized as “alive,” and those who died between waves or in previous waves were categorized as “deceased.” Any participants who left the study between waves or had missing mortality status in 2018 were excluded from the study. The sex, race/ethnicity, and CMV variables that were used in the survey-weighted linear models were used for survival analyses. For ease of interpretation, the hazard ratios were calculated using the equation *e^coefficient^*, where Hazard Ratio > 1 indicates a (|Hazard Ratio - 1|*100)% increase in mortality risk with every unit increase in the biomarker and a Hazard Ratio < 1 indicates a (|Hazard Ratio - 1|*100)% decrease in mortality risk with every unit increase in the biomarker. We estimated the FDR to account for multiple testing.

As a sensitivity analysis, we explored the effects of CMV IgG Antibody (“CMV”) prevalence on mortality. We ran a CPH model with CMV as a potential predictor of all-cause mortality with age as time points and sex and race as covariates. Additionally, we removed CMV from the CPH hazard model (**Figure 1**, Model 2) and applied it to each immunological biomarker using the entire sample. The same process as previously described was used to calculate hazard ratios and correct for false discovery rate. We calculated the percent differences in the hazard ratios from the CPH models adjusting for CMV (Hazard Ratio A) and the hazard ratios for the CPH models without adjusting for CMV (Hazard Ratio B) using the equation 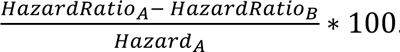.

#### Inferring differences in association sizes attributed to sex

We performed bootstrapping to estimate the influence of sex on ICBs’ association with mortality risk. We applied the same CoxPH models to each immunological biomarker using the entire sample but without sex as a covariate (**Figure 1**, Model 4). We also computed the hazard ratios *e^coefficient^* and applied the FDR method to p-values from these model outputs. Additionally, we calculated the percent differences in the hazard ratios from the CoxPH adjusting for sex group (Hazard Ratio A) and the hazard ratios for the CoxPH models without adjusting for sex (Hazard Ratio B) using the equation 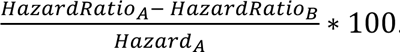 We used the boot() package in R, and calculated the significance of the influence using pnorm().

To complement these analyses, we also investigated whether ICBs affect the association between sex and mortality risk. Bootstrapping analyses were applied to the percent change in hazard ratios for sex after adding each ICB to the CoxPH model. Finally, bootstrapping analyses were conducted to assess the cumulative effect of all ICBs on the hazard ratios for sex.

## RESULTS

**Table 1** presents sample characteristics of the 8,822 participants included in the analysis. The average age at the year of collection was 71 years (sd = 9) **(Figure S1).** 58% were women. Sixty-eight percent of participants were Non-Hispanic White, 18% of participants were Non-Hispanic Black, and 14% of participants were Hispanic (Mexican and Other Hispanic). The mean prevalence of CMV IgG antibodies was 338 U/mL (sd = 439 U/mL). Age and gender distributions were not notably different by racial group, although CMV did differ significantly by racial group (p_white v. black_ = 1.22e-21, p_white v. hispanic_= 2.65e-25). Black and Hispanic groups both had significantly greater CMV IgG antibody levels on average with the mean levels being 472 U/mL (sd = 681 U/mL) for Non-Hispanic Black participants, 442 U/mL (sd = 372 U/mL) for Hispanic participants, and 280 U/mL (sd = 349 U/mL) for Non-Hispanic White participants.

**Table 1:** Participant Demographics. Age, CMV IgG Antibody, and racial group statistics stratified by sex for the dataset used for analyses.*Age in 2016, the year of sample collection. Abbreviations: Cytomegalovirus (CMV), Immunoglobin G (IgG)

| Variable | Female, N = 5,094<sup>†</sup> | Male, N = 3,728<sup>†</sup> |
| --- | --- | --- |
| **Age, years\*** |  |  |
| Minimum | 57 | 57 |
| 25% | 63 | 63 |
| Median | 69 | 69 |
| Mean (SD) | 71 (9) | 71 (9) |
| 75% | 78 | 78 |
| Maximum | 107 | 100 |
| **CMV IgG Antibodies** |  |  |
| Minimum | 0 | 0 |
| 25% | 1 | 0 |
| Median | 293 | 121 |
| Mean (SD) | 391 (494) | 266 (336) |
| 75% | 662 | 438 |
| Maximum | 22,801 | 1,817 |
| **Racial_Group** |  |  |
| Black | 986 (19%) | 597 (16%) |
| Hispanic | 732 (14%) | 545 (15%) |
| White | 3,376 (66%) | 2,586 (69%) |
| <sup>†</sup> n (%) |  |  |

### Differences in ICBs by sex

First, we sought to confirm whether our analysis could identify previously described ICB differences between sexes. Most ICBs were associated with sex. Specifically, 42 out of 53 ICBs were statistically different by sex (−14.153-34.46%) **(Figure 2, Table S1).** 10 of the 48 ICBs differed by more than 10% between males and females **(Figure S2),** including Monocyte count (percent difference -14.1%), CD16- Monocyte Count (−13.9%), and CD8+ T cell percentage (−10.6%) which were lower in females than in males (p values < 0.001). B Cell percentage (percent difference 14.55%), CD4+ Naïve T Cell percentage (14.4), CD4+ T cell Count (17.2%), B Cell Count (15.5%), Naïve B Cell Count (15.5%), CD8+ Naïve T Cell Count (22.9%), CD4+ Naïve T Cell Count (31.7%), and CD8+ Naïve T Cell Percentage (34.4%) were higher in females than in males. None of the five cytokine ICBs were more than 10% different by sex, and TNFR-1 was not significantly different by sex (Figure S3)

**Figure 2:**
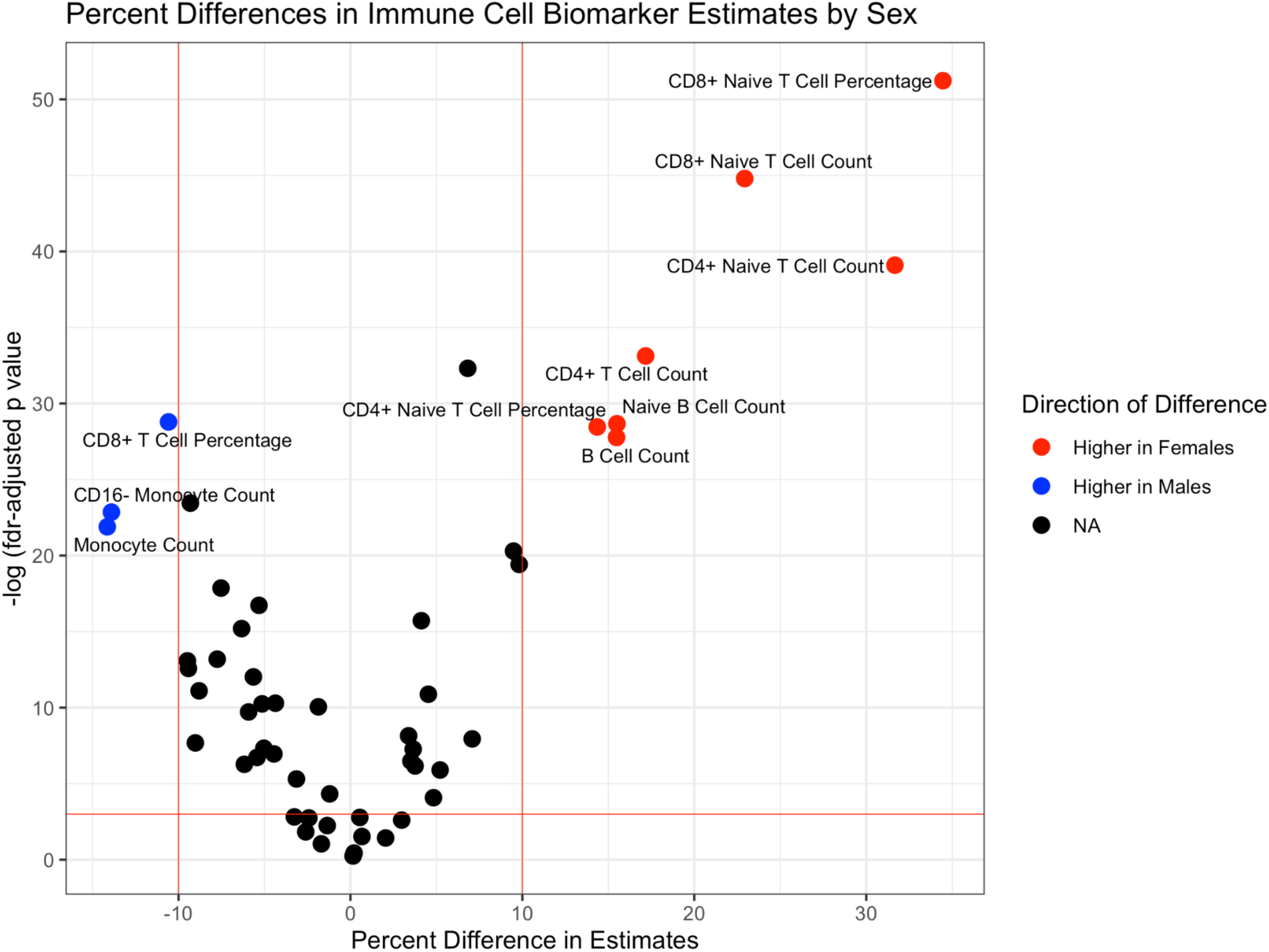
Sex Differences in Immune Cell Biomarkers. This volcano plot highlights statistically and practically significant sex differences in ICBs obtained from a survey-weighted linear model. X-axis: percent difference in biomarker estimates between females and males; >0 = higher in females, <0 = higher in males . Y-axis: -log10 (false discovery rate (FDR)-adjusted p-value), indicating statistical significance. Vertical red lines indicate a magnitude of percent difference >10%. The horizontal red line indicates the threshold for statistical significance (FDR-adjusted p < 0.05). Points are colored by the direction of difference: red indicates higher estimates in females, blue indicates higher estimates in males, and black indicates no significant difference.

### Impact of sex on ICB association with mortality risk

Within the entire sample and not considering sex, twelve ICBs were significantly associated with mortality risk **(Figure 3, Table S2).** Of these twelve, one SD-increase of NK Cell Percentage was associated with decreased mortality risk (HR 0.75, p_adj_ = 0.002), while one SD-increase in CD16- Monocyte count (HR 1.29, p_adj_ < 0.001), Monocyte Count (HR 1.28, p_adj_ < 0.001), NK CD56HI Percentage (HR 1.20, p_adj_ <0.001), Monocyte Percentage (1.19, p =0.03), B Cell Percentage (HR 1.12, p_adj_ = 0.005), CD4+ Effector Memory T Cell Count (HR 1.10, p_adj_ = 0.03), Naive B Cell Count (HR 1.07, p_adj_ < 0.001), and IgD+ Memory B Cell Count (1.07, p_adj_ < 0.001) were associated with increased mortality risk **(Table S2).** Three of the twelve significent ICBs were cytokines; one-SD increases in IL-10 (HR 1.04, p_adj_ = 0.002), IL-1RA (HR 1.15, p_adj_ <0.001), and TNFR-1 (HR 1.22, p_adj_ <0.001) were associated with increased mortality.

**Figure 3:**
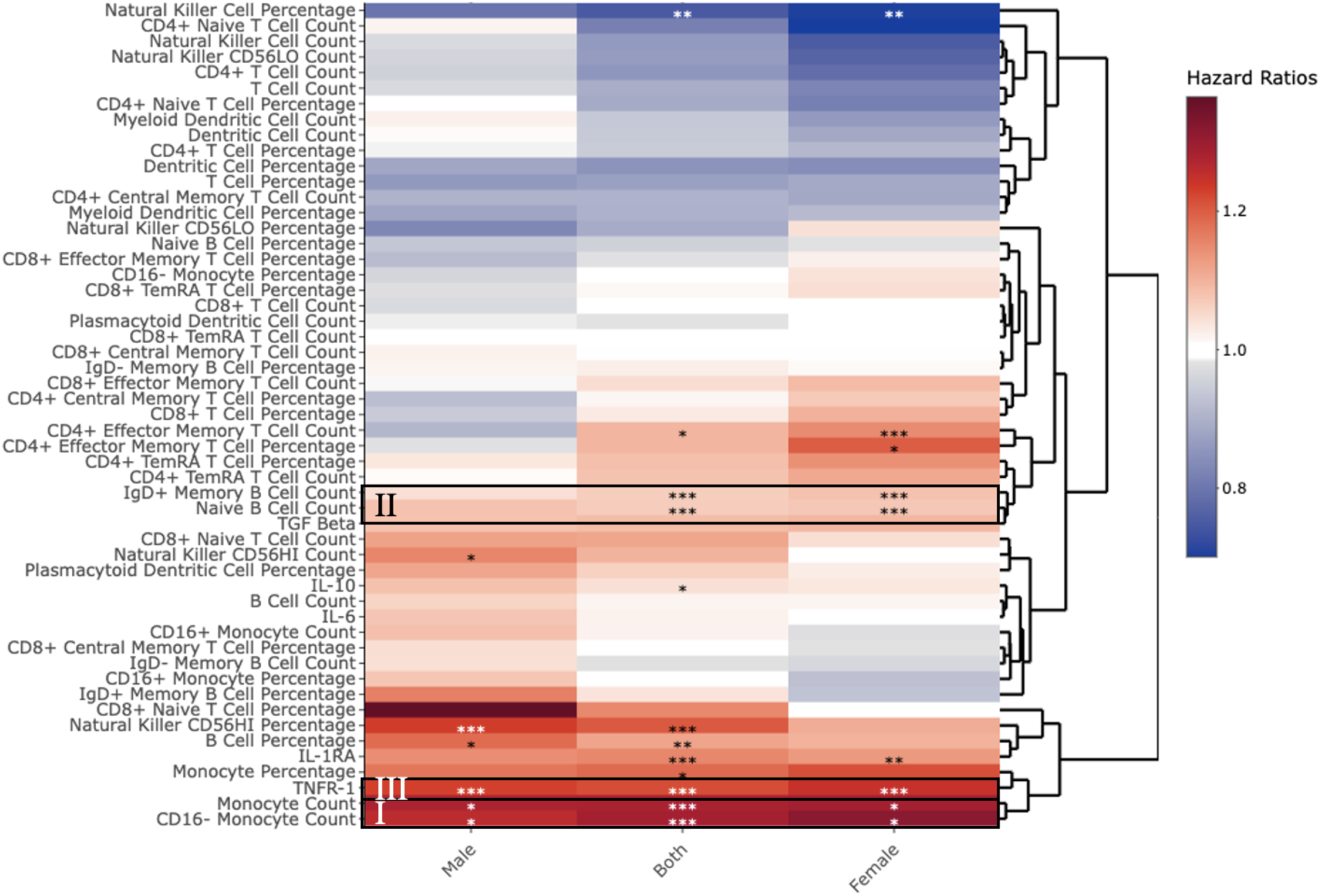
Immune Cell Biomarker Association with Mortality Risk by Sex. This heatmap highlights the similarities and differences in ICB associations with mortality risk in for females, males, and the entire sample (both females and males). ). HR < 1 is represented in blue and indicates a (|HR-1|*100)% decrease in the risk of mortality with every unit increase in the biomarker. HR > 1 is represented in red and indicates a (|HR-1|*100)% increase in the risk of mortality with every unit increase in the biomarker. The clustering of ICBs reflects hierarchical relationships. Columns compare associations for males, females, and the combined cohort. I) Monocyte count and CD16-Monocyte Count are significant predictors of increased mortality risk across all strata. II) Naïve B Cell Count and B Cell Count are predictors of increased mortality risk for females and the combined cohort but not for males. III) TNFR-1 is a significant predictor of increased mortality risk across all strata.

CMV IgG antibody level was not found to be significantly associated with mortality risk (HR 1.07, p = 0.19) **(Table S3).** Furthermore, removing CMV IgG antibody level from the mortality model had a negligible effect (<1.09%) on ICB hazard ratios **(Figure S4, Table S4).**

#### Sex on ICBs association with mortality risk

Next we examined ICBs’ associations with mortality risk within each sex group **(Figure 3, Table S2).** We found that increases in CD16- (Female HR 1.32, p_adj_ = 0.02; Male HR 1.26, p_adj_ = 0.008) and all Monocyte counts (Female HR 1.29, p_adj_ = 0.02; Male HR 1.28, p_adj_ = 0.007) were associated with a higher mortality risk in *both* sexes. TNFR-1 was also associated with higher mortality in both sexes (Female HR 1.25, p_adj_ < 0.001, Male HR 1.23, p_adj_ < 0.001).

In males, increases in NK CD56HI count (HR 1.15, p_adj_ = 0.007) and percentage (HR 1.23, p_adj_ < 0.001) and B Cell percentage (HR 1.18, p = 0.04) was also associated with a significantly increased mortality risk.

In females, CD4+ effector memory T cell count (HR 1.15, p_adj_ < 0.001), CD4+ effector memory T percentage (HR 1.2, p_adj_ < 0.05), IgD+ memory B cell Count (HR 1.08, p_adj_ < 0.001), Naive B cell count (HR 1.07, p_adj_ < 0.001), and IL-1RA (HR 1.13, p_adj_ = 0.003) were also associated with a significantly increased mortality risk. Additionally, increases in NK Cells percentage were only associated with a decreased mortality risk in Females (HR 0.7, p_adj_ <0.001).

Next, we examined how ICB hazard ratios are influenced by sex. We found that there were significant changes in the hazard ratio in seven ICBs **(Figure 4, Table S5).** Most are attenuated by inclusion of sex in the analytic model, which include CD4+ Naïve T Cell Count (4.5%, 95%CI: 1.15-7.81%, p = 0.008), CD4+ T cell Count (3.87%, 95%CI: 1.05-6.74%, p =0.004), CD4+ Naïve T Cell Percentage (3.41%, 95%CI 0.78-6.10%, p = 0.006), TGF-Beta (3.06%, 95%CI 0.87-5.25%, p = 0.003), T Cell Count (2.9%, 95%CI 0.40-5.42%, p = 0.01), CD4+ T Cell Percentage (2.09%, 95%CI 0.26-3.93%, p = 0.01), and TNFR-1 (−0.88%, 95%CI -1.7,-0.07%, p = 0.02). However, others are amplified in their association with mortality on inclusion of sex. These include CD8+ Naïve T Cell Percentage (5.1%, 95%CI 1.3718-8.819.0%, p = 0.0045), TGF-Beta (3.06%, 95%CI 0.87, 5.25, p = 0.003), B Cell Percentage (2.06%, 95%CI 0.56-3.56%, p = 0.003), and Natural Killer CD56LO Count (−1.61%, 95%CI -3.17,-0.04%, p = 0.02).

**Figure 4:**
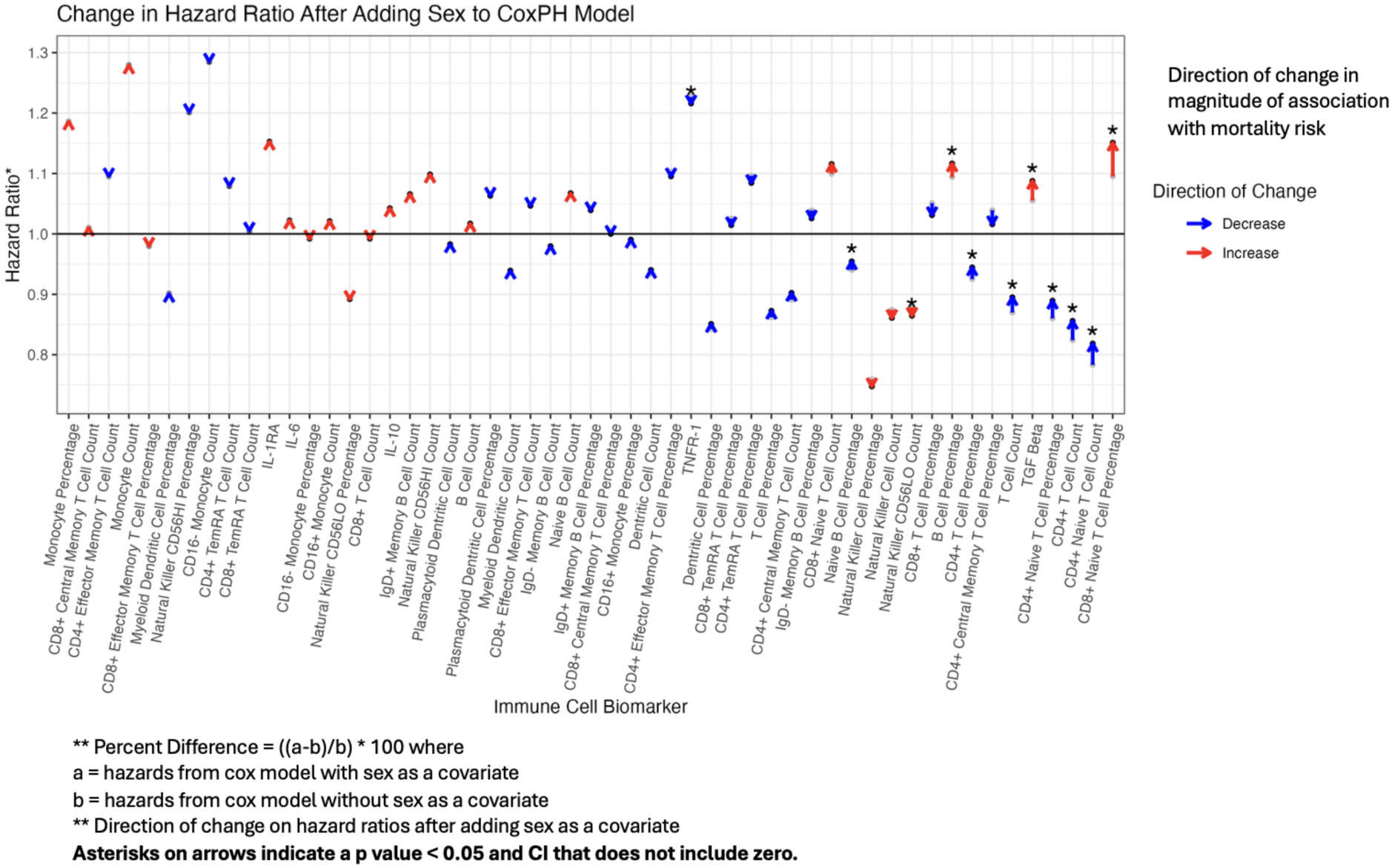
Effect of Sex on ICB Associations with Mortality Risk. This plot highlights the magnitude and significance of the percent change* in hazard ratio after adding sex as a covariate to the CoxPH Model. The horizontal line represents HR = 1 (no association with mortality). Arrows and points below the horizontal line represent ICBs which associated with reduced mortality risk, and those above the horizontal live represent ICBs associated with increased mortality risk. The direction of the arrow indicates the direction of change of the numerical value of the HR, relative to HR =1, after adding sex to the model. The color of the arrow indicates the direction of change on mortality risk; red indicates increased magnitude of association with mortality risk, and blue indicates decreased magnitude of association with mortality risk. Percent change = ((a-b)/b)*100; a = hazards from cox model with sex as a covariate; b = hazards from cox model without sex as a covariate

### Contributions of ICBs to differences in mortality risk by sex

At baseline, females had a significantly different association with mortality risk in comparison to males, with a hazard ratio of 0.66 (p < 0.001), as is well known. In other words, being female is associated with a 34% lower mortality risk in comparison to males.

Adding ICBs to the mortality model did not significantly change the hazard ratio of the sex variable (i.e. the association between sex and mortality risk). We first measured how much the sex hazard ratio changed after adding an individual ICB to the model. We found that the range was between -6.28% and 8.67%; however all changes were non-significant (p > 0.31) **(Table S6).**

We also added all 53 ICBs to examine changes in the mortality risk by sex. The hazard ratio for sex was not significantly effected (HR (− ICBs) 0.66, HR (+ICBs) 0.65, percent change -1.88%, p = 0.44) **(Table S7).**

## DISCUSSION

In our population of older participants in the Health and Retirement Survey, we found that a majority of ICBs differ by sex. In the entire sample, nine ICBs (out of 48) were associated with increased or decreased mortality risk. Importantly, however, the individual and cumulative contributions of ICBs did *not* explain the differences in sex-specific mortality risk in participants of HRS.

Our results on sex differences in T and B cell ICBs correspond with established trends in an aging population. CD4/CD8+ T cell populations exhibit sexual dimorphism across the lifespan, with aged females having increased T cells and decreased CD8+ cell (16,18,34–37). Findings from prior work also suggests that females have more naïve T cells than males after aging (37,38); we correspondingly found that females have increased naïve CD4+ and CD8+ T cell populations and lower CD8 Effector Memory and TemRA T cell percentages. We also found increased B cells in females. In a typical adult population, B cells are known to be higher in females than males (37,39–43). Our corroboration of previous reports emphasizes the stability of T and B cell sex differences in the aged population.

Interestingly, we observed an overlap between the ICBs with significance sex differences in our aging population and the ICBs typically associated with immunosenescence. For example, immune aging is associated with a decrease in naïve and increase in late-differentiated memory T cells (10,12–14). Higher CD8+ T Cells (and consequently an inverted CD4/CD8 ratio) are also “hallmarks” of aging (44–47). This led us to the question of whether the sex differences in ICBs are associated with or analytically explain the mortality differences between males and females.

First, we asked whether ICBs were significantly associated with mortality risk. In the entire sample, including males and females, increases in monocyte and CD16- (Classical) Monocyte count was associated with increased mortality. Monocytes are known to produce increased pro-inflammatory markers, contributing to inflammaging (48) and has thus been previously hypothesized to be a predictor of mortality (49). Our results further emphasize the value of investigating monocytes as a biomarker for all-cause mortality for both sexes.

Interestingly, B cell ICBs showed sex differences in their association with mortality risk. In females only, IgD+ memory B cell count was a predictor of mortality, which corresponds with the trend of increased differentiated leukocytes in immunosenescence. Interestingly, we also found that *increases* in naïve B cell count were associated with increased mortality risk in females while increases in B cell percentage (out of total lymphocytes) was associated with increased mortality risk in males only (15,50,51). We further speculate that in aging individuals, sex-specific regulation of B cell function may contribute to the differences in which B cell populations are associated with mortality risk for each sex.

We also found a range of results associated with sex specificity for NK cells. Increases in NK cell percentage were associated with reduced mortality risk in females and the entire sample (combined males and females). While overall NK cell percentage was also associated with reduced mortality risk in males, the result was non-significant. However, in males, NK CD56HI cells count and percentage (NK cells with high expression of surface marker CD56) were associated with increased mortality risk. Our findings suggest that the change in NK CD56HI cell function contributes to increased mortality risk for males.

While there is a sex-associated axis in T-cell populations at baseline and with aging, which is reflected in our stratified analyses, we found very little differences in sex-specific mortality risk related to T cell populations. The exception was CD4+ effector memory T cell counts, which were found to be associated with increased mortality risk in females but not males. Estrogen receptor (ER) signaling is crucial for regulating CD4+ effector cell (52), which is interesting considering estrogen changes with menopause experienced by aging women. Other ER-regulated functions of CD4+ effector cells include chemokine production (e.g. IFNγ) and ability to activate B cells (53); This process may also be affected by menopause-related hormone changes in aging women and related to the sex-specific B cell associations described earlier, if aging has female-specific effects on the T cells that activate naive B cells.

Overall, our findings are consistent with prior HRS analyses demonstrating that aging-associated immune phenotypes, particularly monocyte and lymphocyte populations, are predictive of mortality (54). Our analyses extend this work by demonstrating that these mortality associations differ by sex, particularly among B-cell and NK-cell subsets, despite the overall modest contribution of ICBs to explaining sex differences in mortality risk. These results highlight the potential of B cell and NK cell subsets as sex-dependent predictors of mortality. We also draw attention to specific ways in which sex-dependent mechanisms acting on these ICBs could be affecting their contribution to mortality. This corroborates other recent work on the HRS finding that immune aging and mortality associations vary across sex and racial/ethnic groups and do so differently depending on the immune biomarker (55).

A main finding of this study is that while different ICBs were associated with mortality risk in males and females, the individual and cumulative contributions of ICBs were non-significant in explaining the mortality risk differences associated with sex. Together with recent studies of immune aging and mortality, our results suggest that compositional immune-cell measurements alone may incompletely capture the biologically relevant dimensions of immune aging linked to survival. For example, a natural next step is to ask how the sex differences in cell counts and percentages and their associations with mortality risk as seen in this study may indicate functional changes in the immune system. These functional changes may help explain sex differences in susceptibility to infection or other conditions (e.g. cardiovascular or autoimmune disease, sepsis) that contribute to all-cause mortality. Moreover, recent work further suggests rather than uniform across populations. Our findings align with this interpretation: although males and females exhibited distinct mortality-associated ICB patterns, these differences did not translate into substantial explanatory power for sex-based mortality disparities overall.

There are a few limitations to consider in this study. The nature of the HRS participant data is that it is only a single time point measurement of the aging population and lack of repeated measures on ICBs so one cannot observe the trajectory of the ICBs through time as it relates to sex and mortality. Second, ICBs were measured using flow cytometry platforms, and more granular or functionally relevant subsets are not captured. We also did not include analysis of ICB ratios (e.g. CD4+/CD8+ T cell ratios) as we intended to conduct an unbiased, broad screening of all available ICBs rather than approach the analysis with key ICBs (or their compositional ratios) in mind. Strengths of this study include approaches to determine the analytical robustness of individual and cumulative contributions of ICBs in explaining sex differences in mortality risk in a representative retiree population of the United States. We also focus on immune cell populations, which are easily measured using peripheral blood, posing it as a potentially clinically-relevant mortality risk biomarker, interrogated in a diverse and representative aging population of the United States.

## CONCLUSION

This study confirms sex differences in key ICBs associated with immunosenescence in an older population. We also identified ICB predictors of mortality risk within our sample. While stratified analysis showed differences in predictive ICBs between males and females, the contribution of sex to ICB associations with mortality risk *and* the contribution of ICBs to sex differences in mortality risk are modest and largely non-significant. Overall, these findings suggest that while there is a sex axis to immune aging detectable in these ICBs, these cell counts and percentage measurements may not be reflective of how immune function contributes to sex differences in mortality risk. Future analyses should consider the mortality associates of functional measures of the ICBs (e.g. monocytes, B cells, or NK cells) that we found to be differently associated with mortality by sex.

## Data Availability

All data produced in the present study are available upon reasonable request to the authors

https://hrs.isr.umich.edu/about

## Abbreviations

HRS: Health and Retirement Study
ICB(s): Immune Cell Biomarker(s)
NK: Natural Killer
CMV: Cytomegalovirus
IL: interleukin
TNF: Tumor Necrosis Factor
VBS: Venous Blood Study
Ig: Immunoglobin
CoxPH: Cox Proportional Hazards Model
TemRA: Terminal Effector Memory
ER: Estrogen Receptor

## Supplementary Figures

**Figure S1:**
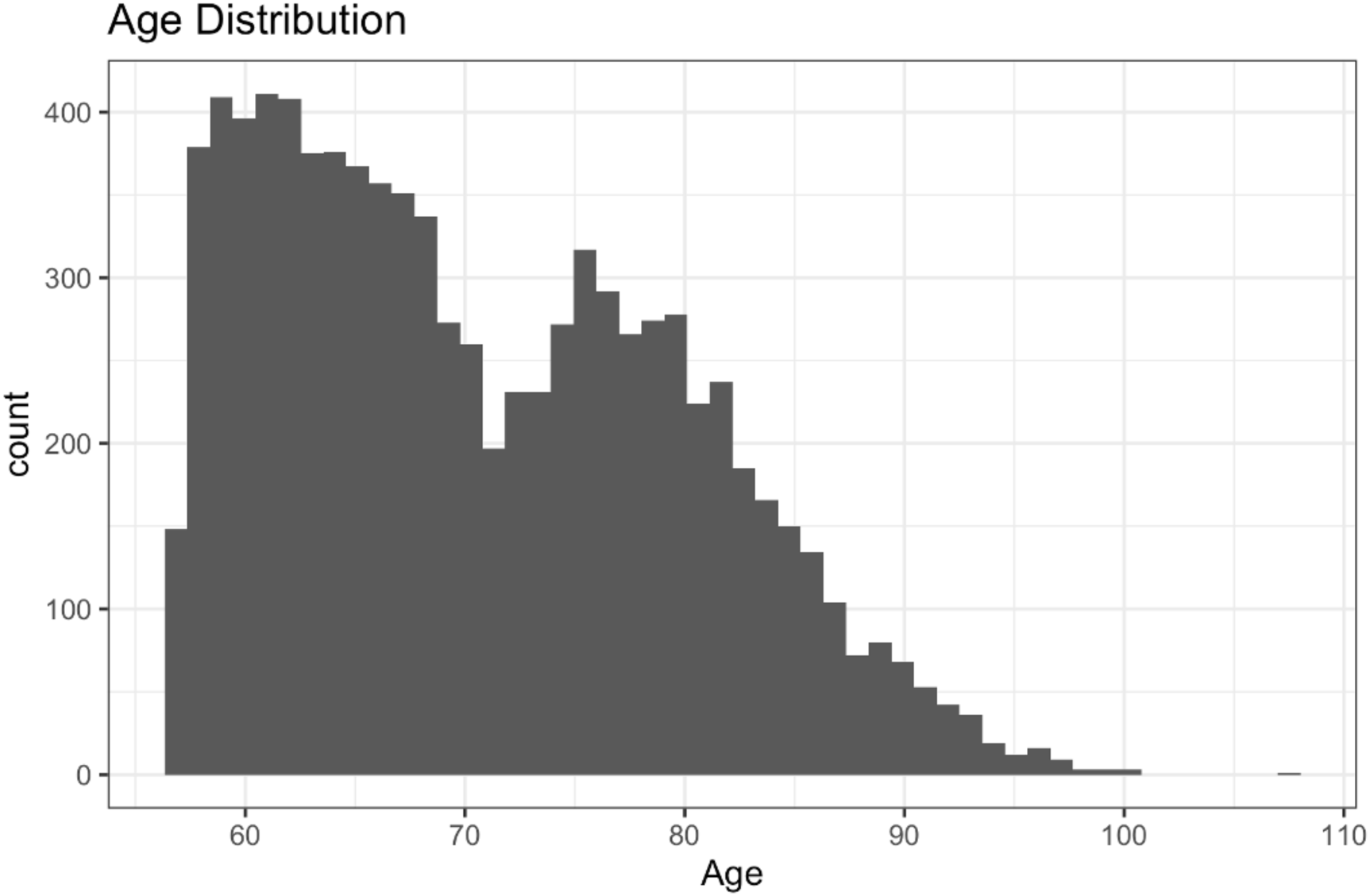
Distribution of Cohort Age. This histogram displays the distribution of the ages of the individuals included in the cohort of HRS participants included in this study. The majority of participants are within ages 60-80 years.

**Figure S2:**
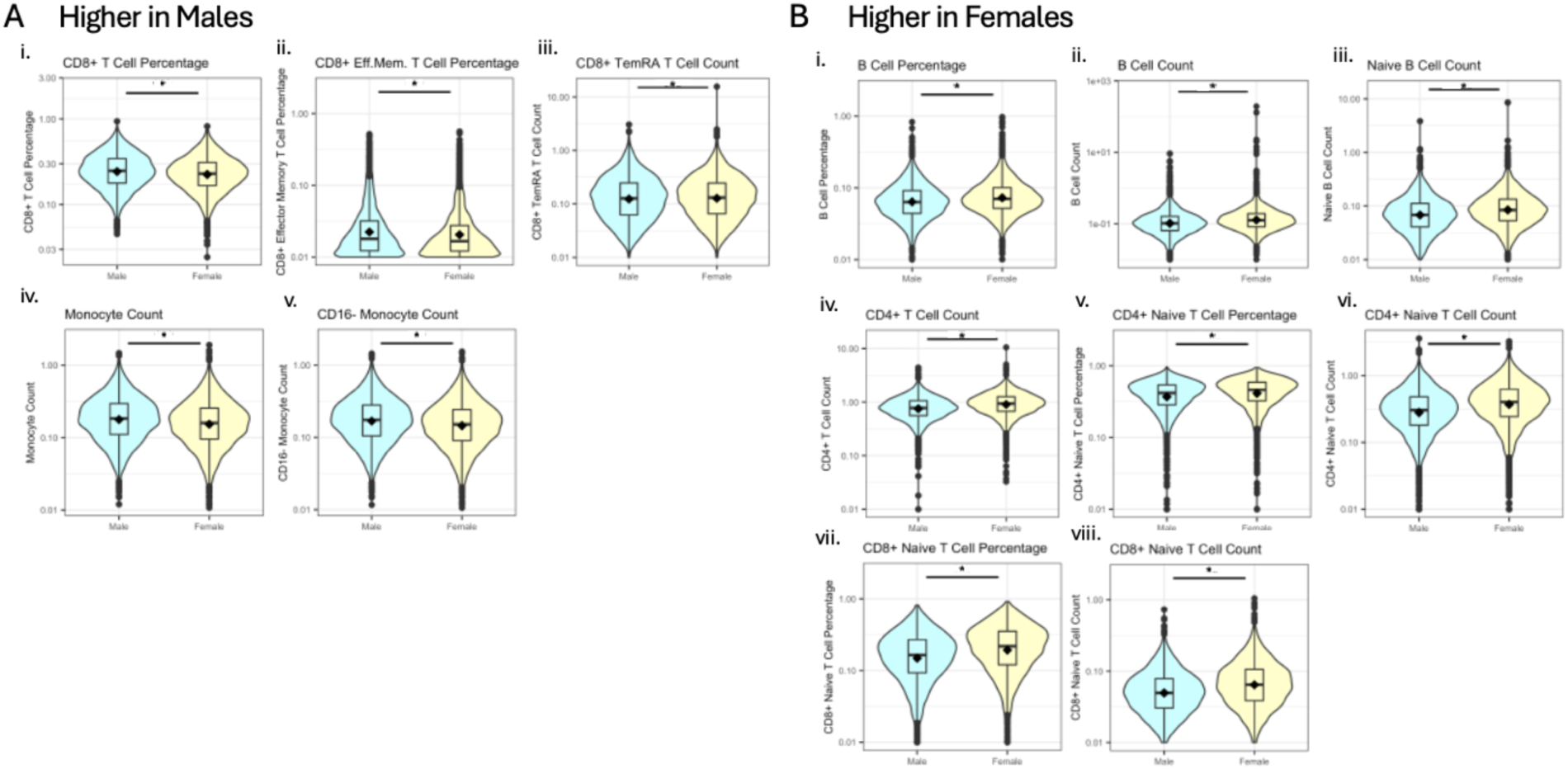
Key ICB Distributions by Sex. These violin plots display the distributions of ICBs (scaled) by sex for ICBs were the percent difference between sexes is greater than 10%. The X-axis is grouped by sex. The Y axis displays the value of the flow cytometry results with a log10 scaled axis. A) Five ICBs were higher in males than females, including CD8+ T Cell Percentage, CD8+ Effector Memory (Eff.Mem.) T Cell Percentage, Cd8+ TemRA T Cell Count, Monocyte Count, and CD16- Monocyte Count. CD8+ T Cell Percentage was defined as a % of total T cells. CD8+ Eff.Mem. T Cell Percentage was defined as a % of total CD8+ T Cells. B) Eight ICBs were higher in females than in males, including B Cell Percentage, B Cell Count, Naïve B Cell Count, CD4+ T Cell Count, CD4+ Naïve T Cell Percentage, CD4+ Naïve T Cell Count, CD8+ Naïve T Cell Percentage, and CD8+ Naïve T Cell Count. B Cell Percentage was defined as a % of total lymphocyte count. CD4+ Naïve T Cell Percentage was defined as a % of total CD4+ T Cells. CD8+ Naïve T Cell Percentage was defined as a % of total CD8+ T Cells.

**Figure S3:**
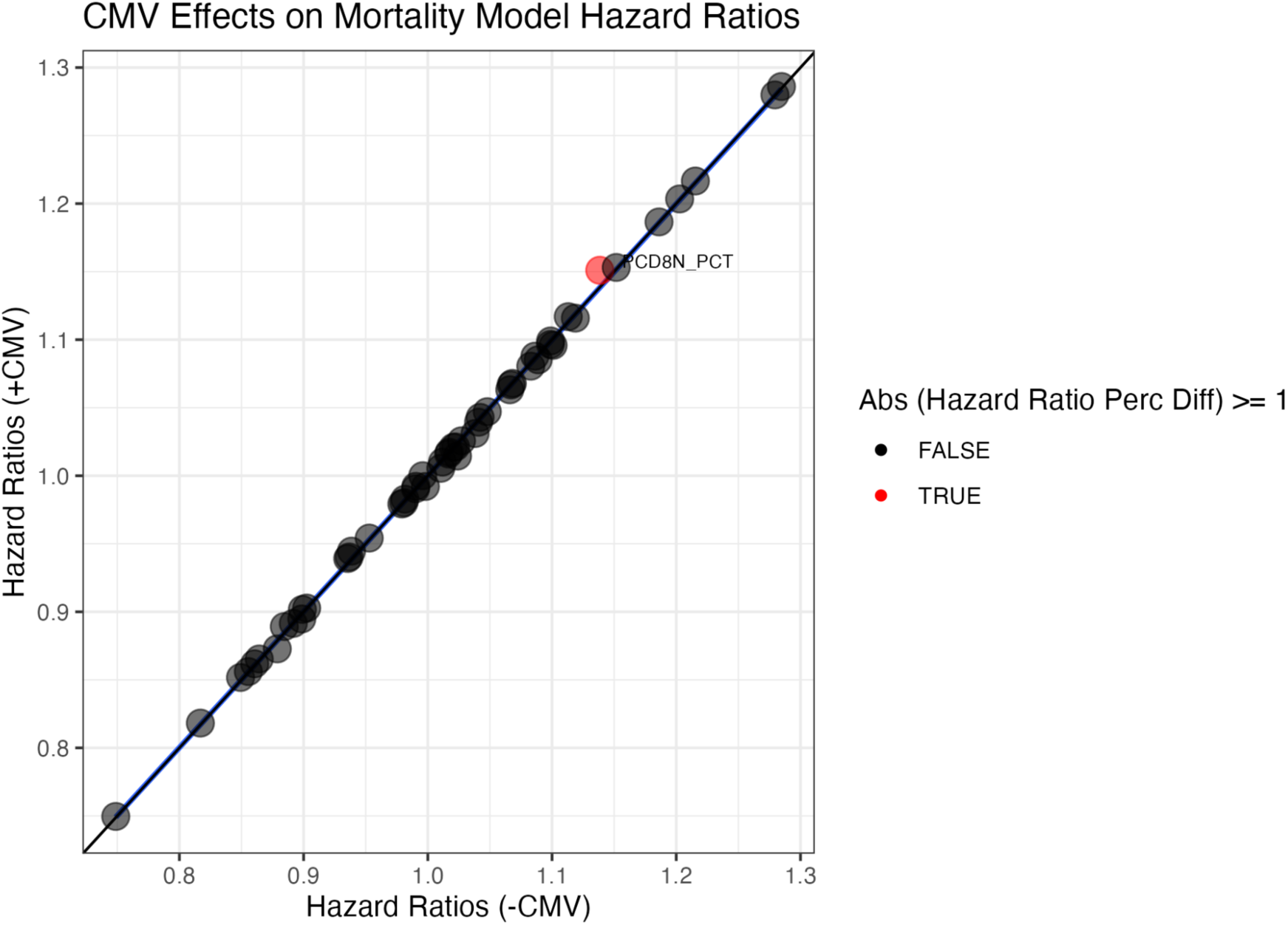
The effects of CMV on ICB associations with mortality. This plot highlights how CMV’s negligible effect on the ICBs associations with mortality risk. The X axis is the hazard ratios from a CoxPH model without CMV as a predictor. The Y Axis is the hazard ratios form a CoxPH model with CMV as a predictor. The black line Y=X represents no change in hazard ratios between the two models. The blue line (barely visibility) is the line of best fit for the points. Points are colored based on the percent differences in the hazard ratios; red represents percent differences greater than 1%, and black represents percent differences less than 1%.

## Declarations

### Ethics approval and consent to participate

The following information is provided in the Institutional Review Board Information for the HRS (https://hrs.isr.umich.edu/sites/default/files/biblio/HRS_IRB_Information%28web%29_08_2018.pdf). Before each interview, participants are provided with a written informed consent information document, read a confidentiality statement, and give oral consent. For the collection of physical measures (including the venous blood collection from which the data in this paper were obtained), written consent form was used to instigate the collection of physical measures at face-to-face interviews starting in 2006. In the 2016 collection, there was also a verbal consent script about willingness to participate and data sharing.

## Consent for publication

Not applicable.

## Availability of data and materials

Any interested researchers are permitted to download and use HRS public use data files at no charge following a brief registration. The RAND HRS Longitudinal File and Tracker File datasets analyzed for this study are available in the HRS repository: https://hrs.isr.umich.edu/data-products The Flow Cytometry dataset analyzed for this study is a HRS Sensitive Health and Genetics Data Products. Sensitive Health Data can be requested by users through the HRS repository through an access agreement.

## Competing interests

The authors do not have competing interests.

## Funding

MY was funded by the Harvard College Research Program. VKN is supported by Harvard Data Science Initiative and diversity supplements through NIEHS R01 ES028802 and NIEHS R01 ES032470. CJP was supported by NIEHS R01 ES032470, NIDDK R01DK137993, NIEHS U24ES03681.

## Authors’ contributions

MY, VKN, and CP conceptualized and designed the study. MY conducted statistical analysis and drafted the manuscript. MY, VKN, AN, and CP provided critical input to analysis strategy, suggested specific analyses, and edited the manuscript.

## Acknowledgements

Not applicable.

## Notes

### Competing Interest Statement

The authors have declared no competing interest.

### Author Declarations

IRB of Harvard University gave ethical approval for this work

